# Investigating Mood and Cognition in People with Multiple Sclerosis: A Prospective Study Protocol

**DOI:** 10.1101/2024.10.02.24314787

**Authors:** Elena C. Cooper, Matthew K. Schindler, Amit Bar-Or, Rachel B. Brandstadter, Monica E. Calkins, Ruben C. Gur, Dina A. Jacobs, Clyde E. Markowitz, Tyler M. Moore, Laura R. Naydovich, Christopher M. Perrone, Kosha Ruparel, Bailey C. Spangler, Scott Troyan, Russell T. Shinohara, Theodore D. Satterthwaite, Erica B. Baller

## Abstract

Multiple sclerosis (MS) is an immune-mediated neurological disorder that affects one million people in the United States. Up to 50% of people with MS experience depression, yet the mechanisms of depression in MS remain under-investigated. Studies of medically healthy participants with depression have described associations between white matter variability and depressive symptoms, but frequently exclude participants with medical comorbidities and thus cannot be extrapolated to people with intracranial diseases. White matter lesions are a key pathologic feature of MS and could disrupt pathways involved in depression symptoms. The purpose of this study is to investigate the impact of brain network disruption on depression using MS as a model. We will obtain structured clinical and cognitive assessments from two hundred fifty participants with MS and prospectively evaluate white matter lesion burden as a predictor of depressive symptoms. Ethics approval was obtained from The University of Pennsylvania Institutional Review Board (Protocol #853883). The results of this study will be presented at scientific meetings and conferences and published in peer-reviewed journals.

**ARTICLE SUMMARY:** *Strengths and Limitations of this Study:*

- We will use MS as a model to study how white matter disease contributes to both the pathophysiology of depression in MS and to general network mechanisms of depression.
- We will leverage research-grade 3-tesla (3T) MRIs acquired as part of routine MS care and maximize scalability by using the Method for Inter-Modal Segmentation Analysis (MIMoSA) for automated white matter lesion segmentation.
- Our study will include participants with medical comorbidities, creating a more representative population and more broadly applicable results.
- We will obtain detailed clinical and cognitive assessments from each participant to evaluate the inter-relationship of mood symptoms, anxiety symptoms, and cognitive deficits, and relate them to white matter disease.
- This is a single-center study.

## INTRODUCTION

Multiple sclerosis (MS) is an immune-mediated neurological disorder that affects one million people in the United States (1,2). It is characterized by demyelinating lesions in the brain and spinal cord due to the immune-mediated attack of oligodendrocytes (3–5). Depression is highly comorbid with MS, with up to 50% of people with MS reporting depression (6,7). Furthermore, 40-60% of people with MS exhibit cognitive dysfunction, and higher levels of depression are associated with greater cognitive impairment (6,8,9). Depression comorbidity in MS contributes to excess morbidity and mortality: it is associated with suicide rates double that of the general population (10–12). Rates of depression in MS are higher than depression comorbidity in other chronic autoimmune diseases, suggesting that the neural pathophysiology of MS may specifically confer increased depression risk (13). Despite the clear overlap between the conditions, the relationship between depression and MS is not understood. The scientific premise of this study is that investigating the relationship between white matter lesions and depression in MS may provide a way to understand the pathophysiology of depression in MS and the network mechanisms of depression more broadly.

Studies of idiopathic depression have described associations between white matter abnormalities and depressive symptoms (14–16). However, research in depression consistently excludes participants with intracranial pathology, and thus results from these studies cannot be extrapolated to people living with MS. Furthermore, MS lesions are heterogeneous – they vary in size, shape, and location (17). Techniques that relate heterogeneous white matter disease to rigorously characterized depression symptoms are vital to fill in this critical knowledge gap.

In contrast to white matter disease in MS, more work has been done relating heterogeneous gray matter disease to the emergence of depression. To overcome limitations in previous studies, Fox and colleagues developed lesion network mapping (LNM) (18), which has shown that strokes in different areas of the brain that cause the same clinical symptoms are part of connected functional networks (19–21). In their most recent work, they extend this paradigm to depression (22–24). The existence of this “depression network” relies on the assumption that network hubs exist on the backbone of structural connectivity. However, few studies have directly evaluated whether white matter lesions in tracts that connect hubs of the depression network similarly produce depression symptoms (25,26).

The purpose of this study is to delineate the relationship between mood and cognition to white matter brain lesions in MS by performing structured assessments of persons undergoing 3T scanning as part of their clinical care. This project will provide critical knowledge regarding how brain network dysfunction contributes to depression, learning from the example of white matter lesions in multiple sclerosis.

### Aim

This study aims to prospectively evaluate white matter disease as a predictor of depressive symptoms in adults with MS. Two hundred fifty participants will be recruited for our prospective cross-sectional sample from the University of Pennsylvania Comprehensive MS Center. Participants will be assessed with structured instruments to measure the severity of depressive symptoms, anxiety symptoms, lifetime psychopathology, and cognitive functioning. This study will assess the relationships among individual variation in white matter lesions and depressive symptoms, while accounting for cognitive functioning and comorbid psychopathology.

### Hypothesis

In a prospective sample, individual variation in depressive symptoms in participants with MS will be associated with white matter lesion burden affecting tracts connecting brain regions in known depression networks.

## METHODS AND ANALYSIS

### Overall Study Design

This is a prospective study that will include data collection. Over the span of the five-year recruitment window (July 1^st^, 2023, to June 30^th^, 2028), we will enroll 250 participants diagnosed with MS who are receiving neuroimaging under the University of Pennsylvania MS 3T clinical MRI protocol for an in-person or virtual research visit (**Figure 1**). Data analyses will take place after the recruitment phase is completed.

**Figure 1.**
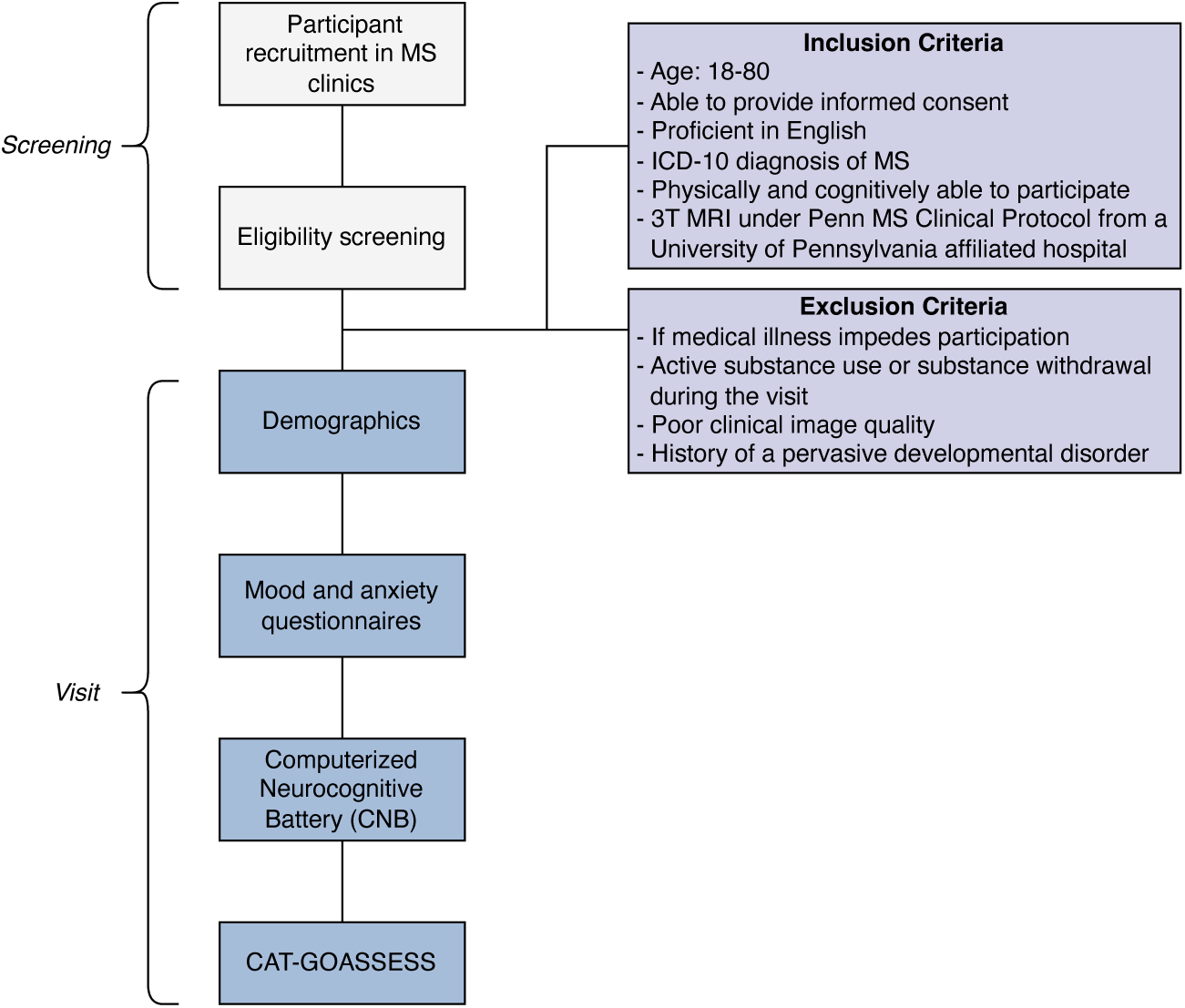
Participants’ Flow Chart. Participants will be screened in MS clinics and then verified for eligibility using the inclusion and exclusion criteria. During the visit, participants will be asked questions about their demographics, answer mood and anxiety questionnaires, complete the Computerized Neurocognitive Battery (CNB) (27), and the CAT-GOASSESS (28). MS, Multiple Sclerosis. CAT-GOASSESS, computerized adaptive version of the GOASSESS.

Participants will undergo a comprehensive neuropsychiatric assessment to assess current and lifetime psychiatric symptoms, as well as cognitive functioning.

#### Symptom Ratings

Participants will complete several symptom rating scales that will measure current levels of depression and anxiety (**Table 1**). This will include: Patient Health Questionnaire Adult (PHQ-9 Adult) (29), Beck Depression Inventory-II (BDI-II) (30), PROMIS Emotional Distress Depression Short (Adult) (31–33), Generalized Anxiety Disorder (GAD-7) (34), Beck Anxiety Inventory (BAI) (35), PROMIS Emotional Distress Anxiety Short (Adult) (31–33) and Hospital Anxiety and Depression Scale (HADS) (36).

**Table 1.** Participant reported outcomes assessed. PROMIS, Patient Reported Outcomes Measurement Information System.

| Scale Administered | Outcome Assessed |
| --- | --- |
| Patient Health Questionnaire Adult (PHQ-9) | Depression |
| Beck Depression Inventory II (BDI-II) | Depression |
| PROMIS Emotional Distress Depression Short (Adult) | Depression |
| Generalized Anxiety Disorder (GAD-7) | Anxiety |
| Beck Anxiety Inventory (BAI) | Anxiety |
| PROMIS Emotional Distress Anxiety Short (Adult) | Anxiety |
| Hospital Anxiety and Depression Scale (HADS) | Anxiety and depression |

#### Assessment of Lifetime Psychopathology

Participants will also complete the computerized adaptive version of the GOASSESS (CAT-GOASSESS) (28), a clinical assessment that provides a dimensional characterization of lifetime psychopathology. Rather than generating clinical diagnoses, which tend to have overlapping symptoms, the CAT-GOASSESS produces transdiagnostic z-scores for 5 domains: mood and anxiety, phobias, externalizing disorders, psychosis, and personality disorders (**Table 2**) (28). Of note, the psychosis domain reflects symptoms of both active and prodromal psychosis, with prodromal questions drawn from the Structured Interview for Prodromal Syndromes (SIPS) (37) and the Prevention through Risk Identification Management and Education screen (PRIME) (38). Personality pathology screening questions were drawn from the Personality Inventory for DSM-5 – Short Form (PID-5-SF) (39).

**Table 2.**
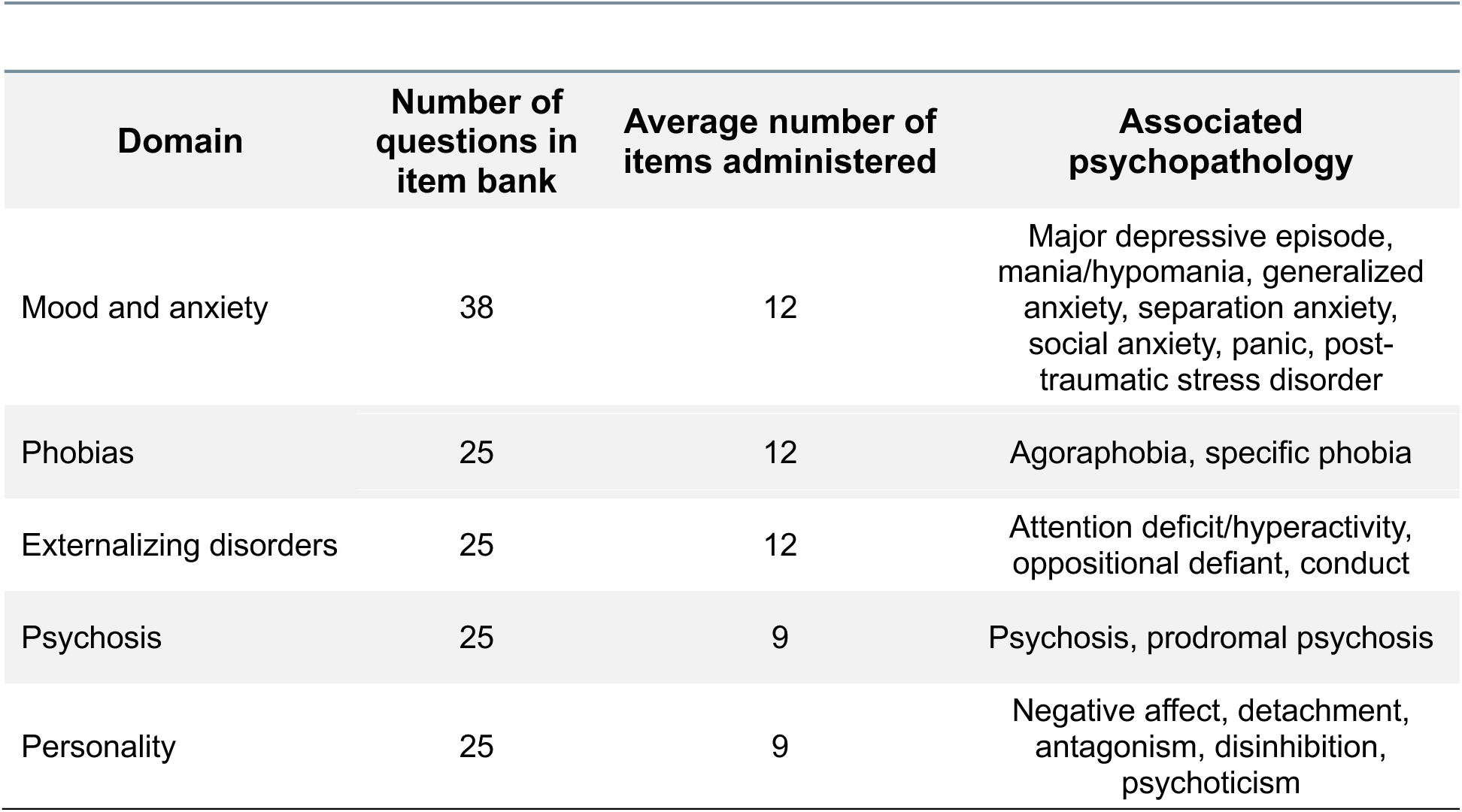
Computerized adaptive version of the GOASSESS (CAT-GOASSESS).

| Domain | Number of questions in item bank | Average number of items administered | Associated psychopathology |
| --- | --- | --- | --- |
| Mood and anxiety | 38 | 12 | Major depressive episode, mania/hypomania, generalized anxiety, separation anxiety, social anxiety, panic, post-traumatic stress disorder |
| Phobias | 25 | 12 | Agoraphobia, specific phobia |
| Externalizing disorders | 25 | 12 | Attention deficit/hyperactivity, oppositional defiant, conduct |
| Psychosis | 25 | 9 | Psychosis, prodromal psychosis |
| Personality | 25 | 9 | Negative affect, detachment, antagonism, disinhibition, psychoticism |

#### Penn Computerized Neurocognitive Battery

Cognition will be assessed with a one-hour computerized neurocognitive battery (CNB) (27) using a version of the CNB that has been developed specifically for MS participants. It will assess thirty-one measures obtained from 17 neurocognitive tests of performance (15 for accuracy, 17 for speed). Domains include executive functioning, episodic memory, social cognition, complex reasoning, and sensorimotor speed. The MS CNB will include the Penn Conditional Exclusion Test, Penn Continuous Performance Task (Number Letter Version – Short), Letter-N-Back Test (2-Back Version), Penn Word Memory Test (plus delayed recall), Penn Face Memory Test (plus delayed recall), Visual Object Learning Test (Short Version + delayed recall), Penn Logical Reasoning Test (Short Version), Penn Matrix Analysis Test (Form A), Variable Short Penn Line Orientation Test (-15 v2.00), Penn Emotion Recognition Test (40 Faces), Measured Emotion Differentiation Test (Form A), Psychomotor Vigilance Test (3 Minute Version), Motor Praxis Test, and the Short Penn Computerized Finger Tapping Test (**Table 3**).

**Table 3.** Computerized Neurocognitive Battery (CNB).

| **Test** | **Domain** | **Accuracy score** | **Speed score** |
| --- | --- | --- | --- |
| Conditional Exclusion Task | Abstraction and mental flexibility | Y | Y |
| Continuous Performance Task | Attention | Y | Y |
| N-Back Test | Working memory | Y | Y |
| Word Memory Test | Verbal memory | Y | Y |
| Word Memory Test - Delayed Version | Verbal memory | Y | Y |
| Face Memory Test | Facial memory | Y | Y |
| Face Memory Test - Delayed Version | Facial memory | Y | Y |
| Visual Object Learning Test | Spatial memory | Y | Y |
| Visual Object Learning Test - Delayed Version | Spatial memory | Y | Y |
| Logical Reasoning Test | Verbal reasoning | Y | Y |
| Matrix Analysis Test | Nonverbal reasoning | Y | Y |
| Line Orientation Test | Spatial reasoning | Y | Y |
| Emotion Recognition Test | Emotion identification | Y | Y |
| Measured Emotion Differentiation Test | Emotion differentiation | Y | Y |
| Psychomotor Vigilance Test | Alertness and vigilance | Y | Y |
| Motor Praxis Test | Sensorimotor speed | N | Y |
| Computerized Finger Tapping Test | Motor speed | N | Y |

#### Medical Record and MRI Imaging Data

Electronic medical record data will be obtained, with participant permission, via the University of Pennsylvania’s Data Analytic Center. We will obtain demographic data, International Classification of Diseases (ICD)-10 diagnoses, medication lists, and depression screens including Patient Health Questionnaires (two question form, PHQ-2; and nine question form, PHQ-9) (29). 3T MRI including 3D T1 magnetization-prepared rapid acquisition gradient echo (MPRAGE) (TR=1.9s, TE=2.48ms, FA=9°, acquisition time=4:18, 176 sagittal slices, resolution=1mm^3^) and 3D T2 FLAIR (TR=5s, TE=398ms, FA=120°, acquisition time=5:02, 160 sagittal slices, resolution=1mm^3^) scans will be acquired as part of routine care and will be transferred directly from radiology to our server.

### Eligibility and Exclusion Criteria

All participants will be 1) males and females aged 18-80, 2) able to provide signed informed consent, 3) proficient in English, 4) physically and cognitively able to participate in a cognitive assessment and complete questionnaires, 5) have an ICD-10 diagnosis of MS from one of the specialists in the University of Pennsylvania Comprehensive MS Center, and 6) receiving routine clinical 3T MRI under the University of Pennsylvania MS protocol from a University of Pennsylvania affiliated hospital.

Subjects will be excluded if 1) medical illness impedes participation, 2) active substance use presents the possibility of acute intoxication or withdrawal, 3) poor clinical image quality, or 4) history of a pervasive developmental disorder.

### Outcome Measures

Outcome measures will include volume of streamlines that intersect lesions at any point in their trajectory (“injured streamlines”) within seventy-seven canonical fascicles as well as disease burden within versus outside the white matter depression network (below). They will be related to the total scores from the symptom ratings, dimensional measures of psychopathology from the CAT-GOASSESS, and cognition from the CNB (**Tables 1-3**).

### Image Processing and Statistical Analysis

#### Constructing White Matter Depression Network

In a series of studies using lesion network mapping (LNM), Fox et al. showed that lesions in the brain that are functionally connected to hubs of the frontoparietal and dorsal attention systems reliably produced depressive symptoms, and that neuromodulation of hubs in this network could alleviate depression (22–24). As we have done previously, we will define the gray matter depression network as areas positively associated with depression (22,25). To construct the white matter depression network, we previously identified fascicles that served as the structural backbone of a functional depression circuit map from Siddiqi et al. (22). We then constructed a binary map by applying a threshold of *t* > 3.09 to identify voxels with a statistically significant positive association between depression symptoms and brain disease or stimulation. The white matter depression network was then created in DSI studio by creating 77 canonical fascicles spanning cortical and subcortical regions from an atlas that was derived from a large sample of high-quality diffusion MRI (31,32). We next calculated the volume (in voxels) that overlapped between the fascicle’s individual fibers, or streamlines, and the binarized functional depression network. Fascicles were ranked by their volume of overlap with the functional depression network. Fascicles with the highest degree of overlap (top 25%, 19 fascicles) were considered to be in the white matter depression network. The resulting white matter depression network included fascicles known to be commonly affected in MS, including the corpus callosum and thalamic projections. However, the superior longitudinal fasciculus and arcuate fasciculus, which support language and complex cognition, also comprised a substantial portion of the white matter network.

#### Lesion Segmentation

The Method for Inter-Modal Segmentation Analysis (MIMoSA) will be used to perform automated lesion segmentation on the MRIs (42). MIMoSA uses local covariance patterns across T1 and T2 FLAIR images within an individual subject to produce a probability map of potential white matter lesions (42). These probability maps will be thresholded and binarized to produce a mask of lesions.

#### Streamline Filtering

We will conduct analyses in template space using tools from DSI Studio, a tractography tool for mapping brain pathways and analyzing tract-related metrics (40,41,43). Specifically, we will project the lesions segmented with MIMoSA onto an expert-vetted, population-averaged atlas of the structural connectome derived from diffusion MRI data (N=842), as reported previously (25,40). This template was achieved by creating a high-resolution template of diffusion patterns averaged across individual subjects and using tractography to generate 550,000 trajectories of representative white matter fascicles annotated by anatomical labels. The trajectories were subsequently clustered and labeled by a team of experienced neuroanatomists in order to conform to prior neuroanatomical knowledge (40). For each individual, the volume of white matter tracts intersecting each lesion will be computed via streamline filtering in DSI studio (40,41,43). This will require identifying whether individual streamlines within a fascicle were impacted (i.e., passed through a lesion) or spared (avoided a lesion). Delineating fascicles in a single diffusion MRI dataset is known to be error-prone, so we will compare spatially normalized lesions to seventy-seven canonical fascicles in template space (44). Individual lesion maps will be normalized to the template space of the canonical fascicles (Montreal Neurological Institute 2009b Asymmetric template) using the T1-weighted-based transform calculated by antsRegistration (45–47). Streamlines intersecting lesions at any point in their trajectory will be considered injured and isolated from the rest of the fascicle. The total volume (in voxels) occupied by injured streamlines will be calculated as the measure of disease burden in the fascicle.

#### Statistical Analysis

For our primary analysis, we will evaluate the relationship of white matter lesion burden within and outside the depression network to depression symptomatology. Secondary analyses will include relating white matter lesion burden to measures of anxiety, cognition, and lifetime psychopathology. Given that MS is a progressive illness that worsens with age, it is vital to account for age effects—which are often nonlinear over the lifespan. Generalized additive models (GAMs, R-package mgcv) (48) with penalized splines will be used to flexibly model both linear and nonlinear aging trajectories while avoiding overfitting by applying a penalty for increasing levels of nonlinearity, which is estimated from the data using restricted maximum likelihood. All analyses will account for multiple comparisons.

### Reproducibility

Data will be managed by DataLad, a free and open-source distributed data management system that allows for the storage of large high-dimensional data and for data analysis to be tracked or undone, thus guaranteeing 100% reproducibility (49). Furthermore, Curation of the Brain Imaging Data Structure (CuBIDS) (50), a Python-based software package that aids users in validating and managing the curation of neuroimaging datasets, will be used to identify heterogeneity in acquisition parameters and ensure all data are in valid BIDS format. All image processing and analysis will be containerized to ensure reproducibility (50).

### Sample Size Power Calculation

To define our sample size target, power calculations were conducted in PASS (NCSS, Version 2022) (51) assuming a 5% type I error rate and using two-sided hypothesis tests. Conservatively assuming 20% of subjects have insufficient data quality (final n=200), we expect to have 80% power to determine associations between depression scores and lesion burden in the depression network with small effect sizes of f^2^=0.04 or larger, adjusting for total lesion volume and age.

### Patient and Public Involvement

Patients and the public were not involved in the design or conduct of this study. Individual results will be communicated to participants who wish to be notified.

## ETHICS AND DISSEMINATION

### Research Ethics Approval

Ethics approval has been obtained from the University of Pennsylvania Institutional Review Board.

### Data Access and Dissemination

All processing pipelines used will be shared via a publicly available software repository on GitHub. As part of our efforts to maximize rigor and reproducibility, all analyses will utilize Jupyter or Rmarkdown notebooks, which will ensure that all interested parties will be able to fully reproduce all analyses. These publicly available notebooks include prose, code, and results. In publication, presentation, or data sharing resulting from this research, the participants will not be identified.

## Data Availability

Due to HIPAA regulations, raw patient data will not be available to the public. All processing pipelines used will be shared via a publicly available software repository on GitHub. All analyses will utilize Jupyter or Rmarkdown notebooks, which will ensure that all interested parties will be able to fully reproduce all analyses. These publicly available notebooks include prose, code, and results.

## AUTHOR STATEMENT

Conceptualization or design of the work, E.C.C., M.K.S., A.B-O., M.E.C., K.R., S.T., R.T.S., T.D.S., E.B.B.; Acquisition, E.C.C., M.K.S., A.B-O., R.B.B., D.A.J., C.E.M., L.R.N., C.M.P., K.R., B.C.S., S.T., R.T.S., T.D.S., E.B.B.; Analysis, E.C.C., R.C.G., T.M.M., R.T.S., T.D.S., E.B.B.; Interpretation, E.C.C., M.K.S., A.B-O., M.E.C., R.C.G., T.M.M., R.T.S., T.D.S., E.B.B.; All authors were responsible for the drafting, revision, and final approval of this manuscript.

